# Diazepam modulates anterior cingulate glutamate levels in people at clinical high-risk for psychosis

**DOI:** 10.1101/2024.12.28.24319730

**Authors:** A. Kiemes, N. R. Livingston, P. B. Lukow, S. Knight, L. A. Jelen, T.J. Reilly, A. Dima, M. A. Nettis, D.J. Lythgoe, C. Casetta, A. Egerton, T. Spencer, A. De Micheli, P. Fusar-Poli, A. A. Grace, S. C. R. Williams, P. McGuire, C. Davies, J. M. Stone, G. Modinos

**Affiliations:** Department of Psychological Medicine, Institute of Psychiatry, Psychology and Neuroscience, King’s College London, London, UK; Institute of Cognitive Neuroscience, University College London, London, UK; Department of Psychiatry, University of Oxford, Oxford, UK; South London and Maudsley NHS Foundation Trust, London, UK; Department of Neuroimaging, School of Neuroscience, Institute of Psychiatry, Psychology and Neuroscience, King’s College London, London, UK; Department of Psychosis Studies, Institute of Psychiatry, Psychology and Neuroscience, King’s College London, London, UK; OASIS Service, South London and the Maudsley NHS Foundation Trust, London, United Kingdom; Department of Brain and Behavioural Sciences, University of Pavia, Pavia, Italy; Department of Psychiatry and Psychotherapy, Ludwig-Maximilian-University Munich, Munich, Germany; Departments of Neuroscience, Psychiatry and Psychology, University of Pittsburgh, Pittsburgh, PA, USA; Brighton and Sussex Medical School, University of Sussex, Brighton, UK; MRC Centre for Neurodevelopmental Disorders, King’s College London, London, UK

**Keywords:** schizophrenia, neuroimaging, benzodiazepines, clinical-high risk for psychosis, glutamate, magnetic resonance spectroscopy

## Abstract

Preclinical evidence suggests that modulating neural excitation through diazepam administration, a positive allosteric modulator of GABA_A_ receptors, can prevent the emergence of behavioural and neurobiological alterations relevant to psychosis in adulthood. Here, we examined this neurochemical mechanism in individuals at clinical high-risk for psychosis (CHRp) in a randomised, double-blind, placebo-controlled crossover study. Twenty-four individuals aged 18-35 were scanned twice using proton magnetic resonance spectroscopy (^1^H-MRS) to measure anterior cingulate cortex (ACC) Glx (glutamate and glutamine) levels, once after a single dose of diazepam (5 mg), and once after placebo. Mixed-effects model analyses revealed that diazepam reduced ACC Glx levels compared to placebo (t(20.8) = −2.14, *p* = 0.04). The effect of diazepam on Glx levels was greater in older CHRp individuals (t(12) = −4.36, *p* = 0.001). These findings suggest that pharmacological modulation of GABA_A_ receptors can alter glutamatergic changes in psychosis.

## Introduction

Individuals at clinical high-risk for psychosis (CHRp) have a 27% risk of developing a psychotic disorder within three years of presentation^1^. However, at present, there are no approved treatments for the prevention of psychosis in this population^2^. Understanding of the neurobiology of psychosis risk is critical for the development of novel preventive treatments, and potentially benefit many young people who may develop poor long term outcomes^3^.

Post-mortem, preclinical, and *in vivo* clinical studies indicate that differences in the interplay between neuronal glutamatergic excitation and GABAergic inhibition play a role in psychosis. Post-mortem studies in schizophrenia have reported reduced expression of GABAergic interneurons in the prefrontal cortex and hippocampus,^4,5^ and reductions in the GABA-synthesising enzyme GAD67^6,7^. Further studies have described reduced mRNA expression of the α1 subunit expression of GABA_A_R in pyramidal neurons in the dorsolateral prefrontal cortex, and elevations in the density of α2 subunits in prefrontal areas^8^. The findings from *in vivo* positron emission tomography (PET) studies using tracers for α1-3 and α5 subunit-containing GABA_A_R (GABA_A_-BZR), the receptors predominantly targeted by benzodiazepines, have been less consistent^9^. One study using [^11^C]Ro15-4513, which has a preferential affinity for the α5 GABA_A_R subunit, reported reduced binding in the hippocampus of antipsychotic-free patients with schizophrenia^10^. However, studies using more general GABA_A_-BZR ligands, such as flumazenil, have not identified differences in GABA_A_-BZR availability in the hippocampus in schizophrenia^9^. Similarly, while one study found an increase of GABA_A_-BZR availability in the anterior cingulate cortex (ACC)^11^, most have not found evidence of altered GABA_A_-BZR availability in the ACC or other cortical regions^9^. Notably, one study focusing on a cohort of CHRp individuals identified reductions in GABA_A_-BZR availability exclusively in the right caudate^12^.

More consistent findings have been evident from *in vivo* proton magnetic resonance spectroscopy (^1^H-MRS) studies of glutamate + glutamine (Glx) levels. In schizophrenia, Glx elevations have been consistently observed in the hippocampus^13,14^. In individuals at CHRp, studies consistently report Glx increases in the medial prefrontal cortex (mPFC), including the ACC, as demonstrated through a meta-analysis^13^. However, hippocampal Glx levels in CHRp individuals show a less consistent pattern. While some studies report increases^15,16^, meta-analyses do not find hippocampal Glx changes^13,14,17^, potentially due to the technical difficulties in Glx ^1^H-MRS imaging in the hippocampus. Glx elevations in schizophrenia have also been correlated with symptom severity: greater mPFC Glx has been associated with greater positive symptom severity, and greater medial temporal lobe Glx has been correlated with greater negative symptom severity^18^.

These findings align with preclinical evidence demonstrating that dysfunction of GABAergic interneurons in the ventral hippocampus can disrupt glutamatergic signalling and broader neural circuits. Studies in the methylazoxymethanol acetate (MAM) developmental rodent model found that GABAergic interneuron dysfunction in the ventral hippocampus leads to hyperactivity and increased glutamatergic outputs from this region^19,20^. Through various downstream multi-synaptic pathways, this mechanism is proposed to drive a hyperdopaminergic state in subcortical regions, which is linked to behavioural analogues of psychotic symptoms^19,21^, mPFC changes related to cognitive symptoms, and alterations in the basolateral amygdala and ACC associated with negative symptoms^19^. Repeated oral administration of diazepam, a positive allosteric modulator of the GABA_A_-BZR site, to MAM-treated rats during puberty prevented the subsequent development of dopamine neuron hyperactivity in the ventral tegmental area^22^, normalised amygdala hyperactivity and anxiety-like behaviour^23^, and reduced the loss of GABAergic inhibitory interneurons in the ventral hippocampus^24^. Independent work on another rodent model involving a genetic knockout of *Erbb4,* a schizophrenia susceptibility gene encoding a tyrosine kinase receptor involved in neuregulin signalling and critical for the maturation and function of fast-spiking interneurons^25^, found that diazepam administration ameliorated schizophrenia-relevant deficits in prepulse inhibition^26^. Our previous research in *Erbb4* conditional mouse mutants using translational *in vivo* neuroimaging revealed increased hippocampal cerebral blood flow and levels of hippocampal glutamine in this model^27^.

The present study aimed to examine the acute, mechanistic effects of diazepam on Glx levels in individuals at CHRp. We selected the ACC as the region of interest because previous research consistently found elevated Glx levels in the ACC and not in the hippocampus in individuals at CHRp^13^. Additionally, diazepam’s GABAergic effects in the hippocampus are hypothesised to modulate glutamatergic projections from the hippocampus to the ACC, leading to downstream reductions in glutamate release and Glx levels in the ACC. We hypothesised that diazepam would increase GABA binding to GABA_A_-BZR on pyramidal neurons, thereby reducing excitatory neuronal activity, glutamate release and glutamate-glutamine cycling, and ACC Glx levels.

### Experimental Procedures

#### Participants

Twenty-four individuals at CHRp aged 18-35 were recruited from the OASIS (Outreach and Support in South London) service^28^. Based on data from previous ^1^H-MRS research at the same scanning site^29^, our sample size was determined via *a priori* power calculation using G*Power version 3.1.9.7^30^ (power = 0.80, alpha = 0.05, two-tailed).

CHRp status was determined using the four positive subscales (unusual thought content (P1), non-bizarre ideas (P2), perceptual abnormalities (P3), disorganised speech (P4)) of the Comprehensive Assessment of At-Risk Mental States (CAARMS)^31^. All individuals were required to exhibit current attenuated psychotic symptoms, determined with a severity or frequency score of ≥3 on P1-P4 of the CAARMS, in the presence or absence of genetic risk and deterioration syndrome or a history of brief limited intermittent psychotic symptoms. Participants fulfilling any of the following criteria were excluded from the study: (1) previous/current exposure to antipsychotic medication at any dosage, (2) diagnosis of a psychotic disorder (3) diagnosis of a neurological disorder, (4) estimated IQ < 70 using the Wechsler Adult Intelligence Scale short version^32^, (5) contraindication to MRI, (6) current exposure to drugs with potential GABAergic or glutamatergic effects (benzodiazepines, anticonvulsants, zopiclone, zolpidem, ketamine, opiates, atomoxetine, memantine, mood stabilisers) other than antidepressants, determined by participant self-reporting and urine drug testing on scanning days and (7) pregnancy/breastfeeding, determined by urine pregnancy test. The study received ethical approval from the London – Bromley Research Ethics Committee (18/LO/0618). All participants provided informed consent in accordance with the Declaration of Helsinki.

#### Procedure

This was a randomised, double-blind, placebo-controlled cross-over design study, by which participants were scanned with MRI twice, once under placebo (50 mg ascorbic acid; Crescent Pharma Ltd, Hampshire, UK)) and once under a single dose of diazepam (5 mg; generic), separated by a minimum three-week washout period. Female participants were mostly scanned with a four-week interval to minimise metabolite fluctuations throughout the hormonal cycle^33,34^. Before the first scanning visit, an assessment visit was conducted to collect sociodemographic information, basic medical history, drug and cigarette use, the shortened Wechsler Adult Intelligence Scale – III^32^ to determine estimated IQ, and clinical measures (CAARMS positive and negative subscales^31^, Hamilton Anxiety^35^ and Depression^36^ scales, Global Functioning Role and Social scales^37^). CAARMS positive and negative composite scores were calculated as sums of subscale severity x subscale frequency (see Supplementary Methods for score calculation).

Prior to scanning, participants were asked to abstain from alcohol, grapefruit-containing products, caffeine for 24 hours and nicotine for 4 hours. To minimise the absorption rate of diazepam^38^, for morning scans (before 13:00), participants were asked to fast from midnight onward, and for afternoon scans (after 13:00), they were asked to eat a light breakfast before 10:00 and fast onwards. Oral diazepam or placebo capsules were administered with 200 ml of water one hour prior to scanning to capture the peak plasma window (1 - 1.5 hours^39^; average peak plasma time 1 hour post administration for 5 mg diazepam^38,40^).

#### Blinding, randomisation and allocation

A diazepam dose of 5 mg was selected on the basis that this produces significant clinical and pharmacokinetic effects without excessive sedation^40^. To maintain blinding of participants, study staff and radiographers, generic diazepam and placebo pills were removed from packaging and placed into an opaque gel capsule (Capsugel, Morristown, NJ) a maximum of one hour prior to prescription collection by the Maudsley hospital outpatient pharmacy. Prescription was collected 30 to 45 minutes prior to dosing time.

A randomisation list was generated by a researcher outside of the study team using a generalised Latin square (Williams Designs) to control for order and first-order carry over effects^41^. Randomisation numbers were allocated one of two possible treatment orders (AB or BA, where A = diazepam and B = placebo). Participants were allocated a sequential randomisation number when the first-scan prescription was requested. Treatment order allocation was kept concealed from the study team at the Maudsley hospital outpatient pharmacy.

#### MRI acquisition

Participants were scanned using a General Electric (Chicago, WI, USA) MR750 3.0 T MR scanner with a 32-channel head coil at the Centre for Neuroimaging Sciences, Institute of Psychiatry, Psychology and Neuroscience, King’s College London. A T1-weighted Inversion Recovery Spoiled Gradient Echo (IR-SPGR) sequence (TE/TI/TR = 3.02/400/7.31 ms; flip angle = 11°; matrix = 256 x 256; FoV = 270; in-plane resolution = 1.05 x 1.05 mm²; slice thickness = 1.2 mm; 196 slices) was acquired for ^1^H-MRS voxel placement and voxel tissue content calculation.

^1^H-MRS spectra were acquired from an ACC voxel (20 x 20 x 20 mm^3^), placed according to previous studies^29,42^, using point-resolved spectroscopy (PRESS; TE/TR = 30/3000 ms; flip angle = 90°; 96 transients) (Figure 1). Briefly, the centre of the voxel was placed along the sagittal midline, 16 mm above the anterior portion of the genu of the corpus callosum, 90° to the anterior commissure to posterior commissure line. Water suppression was performed with the standard GE PROBE (proton brain examination) sequence, which utilises a standard chemically selective suppression (CHESS) water suppression routine. For each spectrum, unsuppressed water reference spectra were acquired (16 transients) to perform eddy current correction and water scaling during spectral analysis. Shimming and water suppression were optimised and by performing auto-prescan twice before each scan.

**Figure 1.**
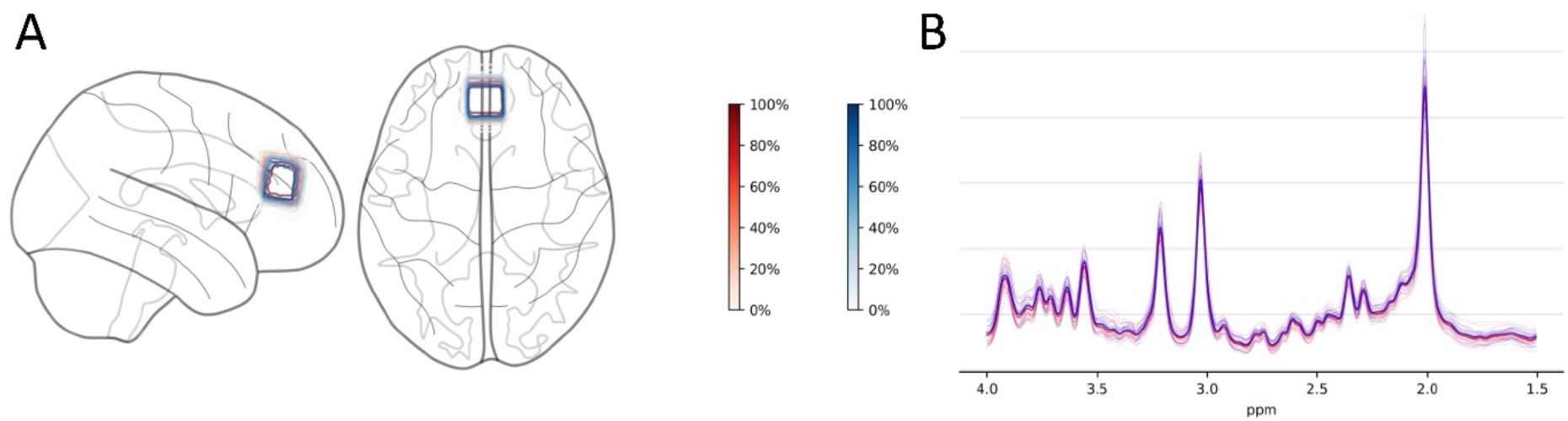
^1^H-MRS voxel placement and spectra overlap. (A) Normalised voxel placement for individual participants and conditions in the anterior cingulate cortex. Shading of contour lines on the glass brain and the colour scales indicate the percentage of voxel overlap across participants within a condition, with darker red areas showing greater overlap and lighter shades indicating lower overlap. (B) Fitted spectra of individual participants and conditions and average spectra per condition in the anterior cingulate cortex. Red, placebo condition; blue, diazepam condition. Figure generated using MRS-voxel-plot^43^ with nilearn^44^. ppm, parts per million.

Spectra were pre-processed using FID-A toolkit^45^. This included multi-coil combination, removal of bad averages, and frequency and phase drift correction. Subsequently, spectra were analysed in LCModel 6.3-1N. Water-suppressed spectra were fitted to a standard basis set included in LCModel, including 17 metabolites (L-alanine, aspartate, creatine (Cr), phosphocreatine (PCr), γ-aminobutyric acid, glucose, glutamine, glutamate (Glu), glutathione, glycero-phosphocholine, phosphocholine, myo-Inositol (mI), L-lactate, N-acetylaspartate (NAA), N-acetylaspartylglutamate, scyllo-inositol, and taurine) and was acquired using PRESS with the same echo time (30 ms) and at the same field strength (3T). To account for random variation in data fitting to basis set, a bootstrap analysis was performed. Linear combination modelling within LCModel was performed iteratively (50 modelling replicates). Due to non-normal distribution of replicate linear combination modelling output, median values were used for subsequent statistical analysis (sensitivity analysis using mean values was performed and are found in our supplementary materials and results).

Initial quality control of spectra involved assessment of signal-to-noise ratio (S/N) and spectral linewidth (full width at half-maximum; FWHM), and visual inspection. Spectra of FWHM > 2 standard deviations above the mean and S/N < 2 standard deviations above the mean^46^, and data points with Cramér Rao Lower Bound (CRLB) minimum variance >20% were excluded. To correct water-scaled metabolite values for voxel tissue composition, segmentation was performed using Gannet 3.1 (Matlab 9.5.0; SPM 12; FSL 5.0.11) to attain the fraction of cerebrospinal fluid (CSF), grey matter (GM) and white matter (WM) per voxel per individual. Metabolites (M) were corrected for CSF according to the following formula: 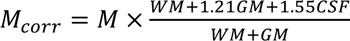. This formula assumes a water tissue concentration of 55,556 mmol/L in CSF, 35,880 mmol/L in WM and 43,300 mmol/L in GM^47^. Concentration of metabolites is expressed in institutional units (i.u). The MRSinMRS checklist^48^ is reported in supplementary materials (Table S1).

#### Statistical analysis

Statistical analyses were performed using R (v4.2.2; https://www.r-project.org/). A significance threshold of *p* < 0.05 was set for all analyses. Firstly, all within-group differences in data quality (FWHM, S/N) and voxel tissue composition (GM, WM, CSF) between the placebo and diazepam conditions were assessed via paired t-tests. Secondly, we analysed changes in our *a priori* metabolite of interest (Glx) between the placebo and diazepam conditions using a weighted linear mixed effects model (*lme4* package). Our primary mixed effects model included participant ID as a random factor and treatment as fixed effect within-subject factor. As supplementary analyses, we performed two further models. To account for potential confounding treatment order, interscan interval and data quality effects, treatment order and days between scans were added as covariates of no interest, and data quality (1-FWHM) as weight factor. A second supplementary model investigated potential confounding effects of age, sex, current antidepressant treatment status^49^, and cigarette use (cigarettes/day)^50–52^ by including these variables as covariates of no interest on top of the previous mixed effects model analysis. No outliers were detected using Grubbs outlier detection.

Exploratory regression analyses investigated whether baseline clinical characteristics (CAARMS positive or negative composite scores) were related to Glx change (diazepam minus placebo) in the ACC. Both CAARMS subscale composite scores were entered into one multiple regression model. Potential confounding effects of age, sex^53^, current antidepressant treatment status^54^, and cigarette use^50^ on these associations were explored by adding these variables as covariates of no interest in a second model. Significance was set at *p* < 0.05, and multiple comparisons corrections were not performed as these multiple regression models were exploratory.

## Results

### Sample characteristics

A total of 24 individuals at CHRp with a mean age ± standard deviation (SD) of 24.1 ± 4.8 years were scanned twice. Demographic and clinical characteristics of the sample are presented in Table 1.

**Table 1.** Demographic and clinical characteristics.

|  | Variable | CHRp individuals (N = 24) |
| --- | --- | --- |
| **Demographics** | Age, mean ± SD years | $24.1 \pm 4.8$ |
|  | Age range, years | 18 - 32 |
|  | Sex, *n* female/male | 15/9 |
|  | Handedness, *n* right/left | 23/1 |
|  | Ethnicity, *n* White/Black/Asian/Mixed | 12/6/2/4 |
| | IQ, mean ± SD | $97.7 \pm 21.6$ |
| **Clinical** | CAARMS positive composite, mean ± SD | $46.4 \pm 13.0$ |
| | CAARMS negative composite (*n*=22), mean ± SD | $29.4 \pm 24.2$ |
| | HAM-A (*n*=22), mean ± SD | $17.1 \pm 8.7$ |
| | HAM-D (*n*=21), mean ± SD | $13.9 \pm 6.9$ |
| | GF:S current, mean ± SD | $6.4 \pm 1.5$ |
| | GF:R current, mean ± SD | $6.1 \pm 1.8$ |
|  | Current antidepressant medication, *n* yes/no | 9/15 |
| **Substance use** | Current cigarette use, *n* yes/no | 8/16 |
| | Cigarettes/day in smokers, mean ± SD | $1.8 \pm 1.6$ |
|  | Current cannabis use, *n* yes/no | 7/17 |

*IQ, estimated intelligence quotient using the shortened Wechsler Adult Intelligence Scale; CAARMS positive composite, composite score (severity x frequency) of positive subscales (P1-4) on the Comprehensive Assessment of At Risk Mental States; CAARMS negative composite, composite score (severity x frequency) of negative subscales (4.1-4.3) on the Comprehensive Assessment of At Risk Mental States; HAM-A, Hamilton Anxiety Rating Scale; HAM-D, Hamilton Depression Rating Scale; GF:S, Global Functioning Social Scale; GF:R, Global Functioning Role Scale*

Participants were scanned with a mean ± SD of 30 ± 8 days interscan interval (female participants: M ± SD = 28 ± 4 days; male participants: M ± SD = 33 ± 11 days). The range and mean ± SD of time from dosing to ^1^H-MRS scanning was 52-97 min and 71 ± 9 min. All participants completed both ACC ^1^H-MRS scans. Data quality parameters are presented in Table 2. No significant differences in spectral quality or voxel tissue composition between the diazepam and placebo conditions were observed. Due to spectral artefacts and quality control failure, two datapoints from the diazepam condition and one datapoint from the placebo condition were excluded. These excluded datapoints were from three different participants, who were not removed from the overall analysis due to incomplete data sets. No further data was excluded based on Cramér Rao Lower Bound (CRLB) minimum variance (>20%).

**Table 2.** Water-scaled glx levels and Spectral quality data.

|  | **Anterior cingulate cortex ($n = 24$)** |  |  |
| --- | --- | --- | --- |
|  | Placebo | Diazepam | Statistics |
| **Glx levels, i.u.** | $20.17 \pm 2.49$ | $18.99 \pm 1.97$ | $t(20.8) = -2.14, p = 0.04$ |
| **Glx CRLB, %** | $5.39 \pm 0.68$ | $5.66 \pm 0.67$ | $t(23) = -1.41, p = 0.17$ |
| **FWHM, ppm** | $0.03 \pm 0.01$ | $0.03 \pm 0.01$ | $t(23) = 0.01, p = 0.99$ |
| **S/N** | $33.46 \pm 4.92$ | $33.19 \pm 6.84$ | $t(23) = 0.22, p = 0.82$ |
| **GM** | $0.66 \pm 0.05$ | $0.66 \pm 0.05$ | $t(23) = -0.17, p = 0.86$ |
| **WM** | $0.09 \pm 0.03$ | $0.09 \pm 0.03$ | $t(23) = 1.05, p = 0.30$ |
| **CSF** | $0.25 \pm 0.04$ | $0.26 \pm 0.04$ | $t(23) = -0.52, p = 0.60$ |

*Mean ± SD are displayed. Glx, glutamate + glutamine; i.u., institutional units; CRLB, Cramér-Rao Lower Bounds; FWHM, full width half maximum; ppm, parts per million; S/N, signal-to-noise ratio; GM, grey matter fraction; WM, white matter fraction; CSF, cerebrospinal fluid fraction*

### Effect of diazepam on ACC Glx levels

In our primary analysis, ACC Glx levels were significantly lower under the diazepam condition (M ± SD = 18.99 ± 1.97) compared to the placebo condition (M ± SD = 20.17 ± 2.49; t(20.8) = −2.14, *p* = 0.04, d = 0.47; Table 2, Figure 2). Similarly, ACC Glx levels were reduced under diazepam when accounting for treatment order, days between scans and quality parameters (first supplementary model: t(20.7) = −2.11, *p* = 0.047, d = 0.46). Adding age, sex, antidepressant treatment, and cigarette use to the model (second supplementary model) rendered the observed treatment condition effect non-significant (t(20.9) = −2.04, *p* = 0.055, d = 0.45). All controlling variables had no significant effect in this model (see Table S2 for contribution of covariates in supplementary models).

**Figure 2.**
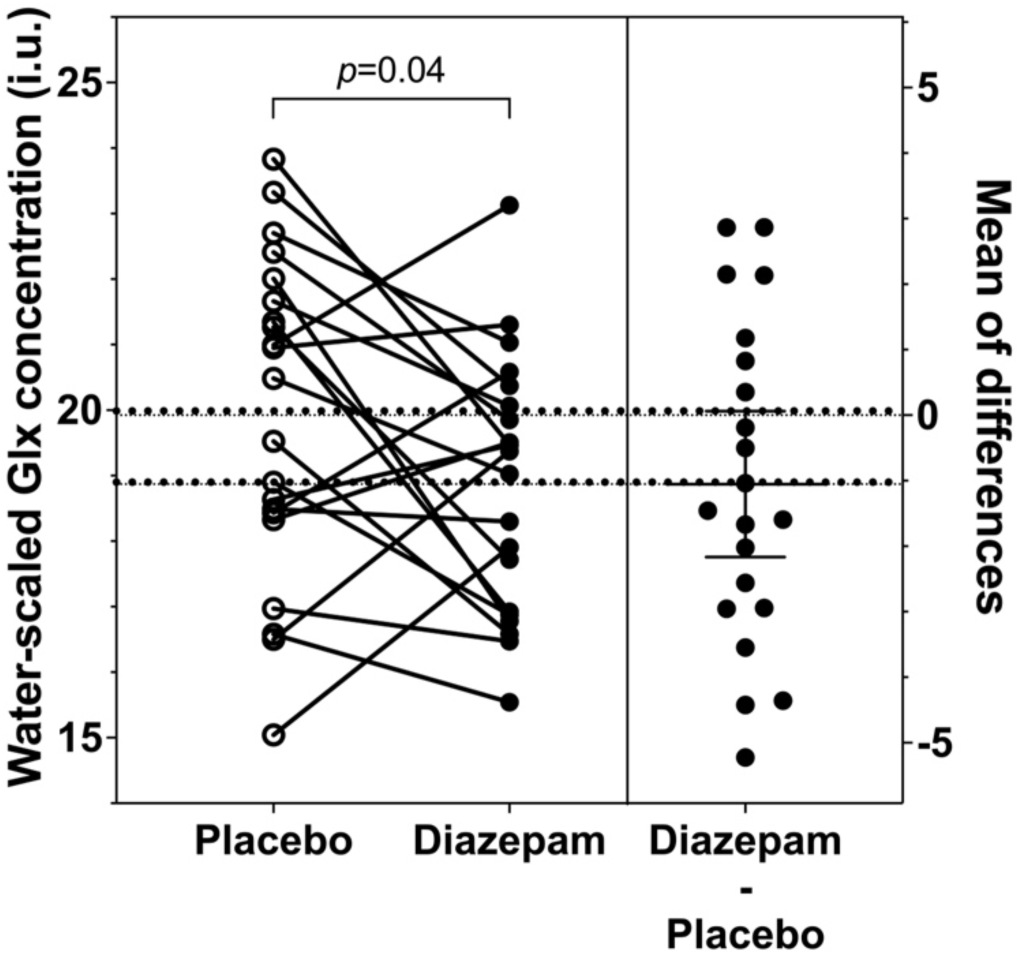
Glx levels in the anterior cingulate cortex during placebo and diazepam administration. Glx levels in the anterior cingulate cortex in individuals at clinical high-risk for psychosis were reduced by acute diazepam administration. Individual Glx levels under the placebo and diazepam conditions are presented on the left and individual diazepam-induced Glx change (diazepam – placebo) with mean and 95% CI are presented on the right. Incomplete datasets were removed for illustrative purposes. Glx, glutamate + glutamine.

### Exploratory analyses

Baseline CAARMS positive or negative composite scores were not associated with the diazepam-induced change in ACC Glx compared to placebo (model summary: *R^2^* = 0.05, *F*(2,16) = 0.44, *p* = 0.65). Adding sex, age, antidepressant treatment and cigarette use as covariates rendered the regression model significant (model summary: *R*^2^ = 0.65, *F*(6,12) = 3.73, *p* = 0.02; Figure 3). This was driven by a significant negative association between diazepam-induced change in ACC Glx levels and age (t(12) = −4.36, *p* = 0.001, d = 2.52). Further regressor statistics are reported in the supplementary results (Table S3).

**Figure 3.**
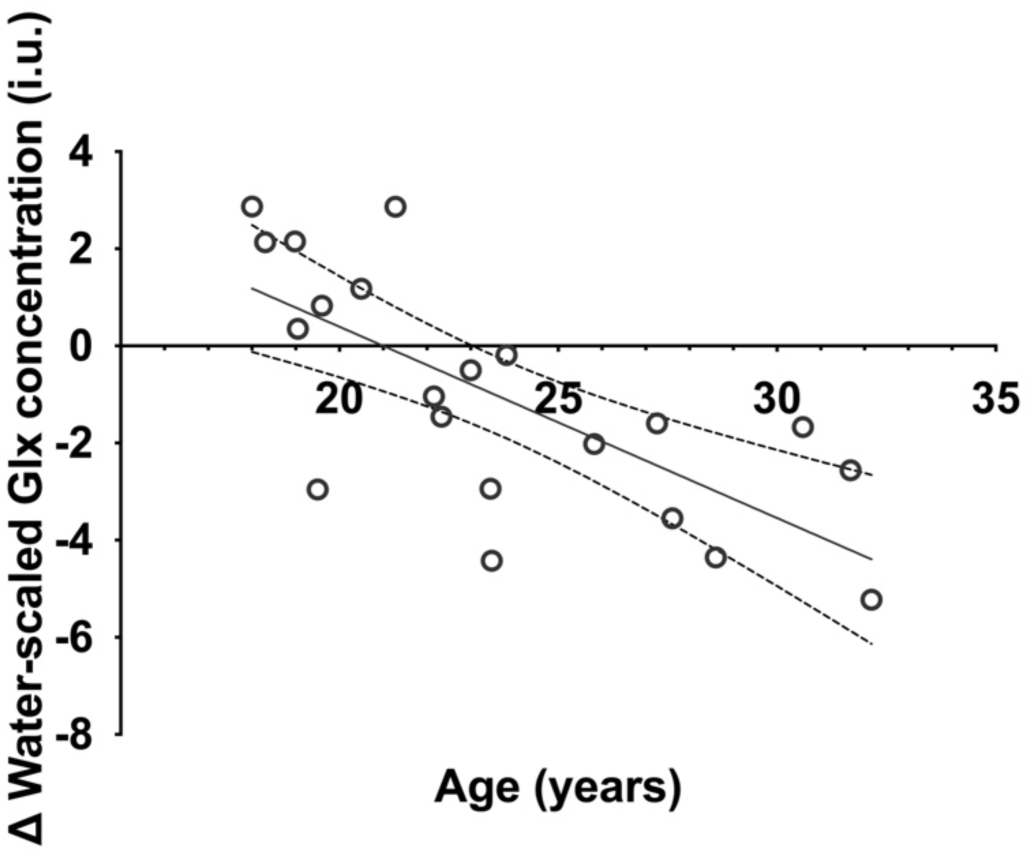
Change in anterior cingulate cortex Glx over age of participants. Multiple regression model demonstrated a negative association with Glx change and age (diazepam Glx levels minus placebo Glx levels; t(12) = −4.36, *p* = 0.001). Interindividual diazepam-induced ACC Glx change was negatively associated with age of participants. Glx, glutamate + glutamine.

## Discussion

Our study revealed that an acute diazepam challenge modulated ACC Glx levels in CHRp individuals, which were significantly reduced compared to placebo. There was an effect of age on these findings, by which the greatest diazepam-induced reductions in ACC Glx levels were observed in older CHRp individuals. These results provide preliminary support for the potential use of GABA-enhancing compounds as a therapeutic strategy to regulate brain excitation in early psychosis.

Previous ^1^H-MRS studies in individuals at CHRp and in patients with schizophrenia have demonstrated alterations in Glx (glutamate + glutamine) and glutamine in the mPFC, including the ACC^13^. In line with our hypothesis, diazepam reduced ACC Glx levels versus placebo in CHRp individuals. This suggests that elevated cortical glutamatergic function in this population may be reduced by positive allosteric modulation of GABA_A_-BZR. Mechanistically, diazepam-enhanced inhibition may reduce glutamate release and glutamate-glutamine cycling^55,56^, reflected as decreased Glx levels measured by ^1^H-MRS. Although glutamine is estimated to make up approximately 10% of the ^1^H-MRS Glx signal^57^, glutamine may be driving the observed ^1^H-MRS Glx changes, as it is primarily dedicated to neurotransmitter synthesis (≈70-90%)^58^, specifically that of glutamate, and to a much lesser extent GABA^59^. Future studies may tackle this question by ^1^H-MRS sampling in CHRp groups using higher-field (>4 T) scanners, where the glutamate and glutamine signal can be separated more reliably^57^. For example, in a preclinical study using a 9.4 T scanner, we identified elevations in glutamine, but not glutamate, in *Erbb4* conditional mouse mutants, a genetic mouse model relevant to schizophrenia^27^. Overall, together with our recent finding that a single dose of diazepam significantly normalised hippocampal hyperperfusion in the same sample of CHRp individuals^60^, this evidence provides mechanistic support for the use of GABA-enhancing compounds to modulate neuroimaging-based phenotypes in individuals at CHRp, highlighting their potential as therapeutic strategy to mitigate neurobiological changes in this patient group. These findings also align with previous preclinical work demonstrating that diazepam has a therapeutic effect on schizophrenia-relevant behavioural and electrophysiological phenotypes in genetic and neurodevelopmental rodent models^22–24,26^. However, diazepam as well as other benzodiazepines may produce an unfavourable side effect profile due to its non-specificity to general GABA_A_-BZR binding, causing drowsiness, fatigue and confusion^39^. More importantly, prolonged use can result in dependence and receptor composition and function^61^. However, more specific positive allosteric modulators of GABA_A_R avoid the sedation and dependence putatively mediated by effects on the α1 GABA_A_R subunit^62^. In MAM rats, positive allosteric modulation of the α5GABA_A_R^63^ and overexpression of the α5 subunit normalised hippocampal hyperactivity in adult rodents^64^. Thus, compounds with higher GABA_A_R specificity, such as to the a5 GABA_A_R subunit, may have similar efficacy in humans without undesirable side effects.

Exploratory analysis suggested that diazepam-induced reductions in ACC Glx were not significantly associated with baseline symptom severity. However, we observed that age was a significant contributor to the model and was negatively associated with change in ACC Glx, such that younger participants exhibited smaller reduction or even increases in Glx under diazepam and older participants showed greatest Glx reductions under diazepam compared to placebo. A previous mega-analysis^65^ found that variability in Glx levels in patients with schizophrenia compared to healthy volunteers in the mPFC (including the ACC) increased with older age, suggesting greater dysfunctional regulation of glutamate levels with age. Due to the role of GABAergic transmission in the tight regulation of excitatory signalling^66^, diazepam as a GABA-enhancing compound may have greater efficacy in more dysregulated states. Together, these findings suggest that GABA-enhancing compounds may induce greater modulation of brain excitatory metabolites in older individuals. As our sample was not originally statistically powered for these correlational analyses, these findings warrant replication in larger samples. However, this preliminary finding is consistent with an age-related decline in GABA inhibition^67,68^, which may result in greater efficacy of benzodiazepine^69^. In contrast, preclinical studies using MAM-treated rats have demonstrated that chronic peripubertal diazepam administration prevented the development of schizophrenia-like phenotypes, suggesting that a chronic treatment paradigm may be required to achieve effects in younger individuals.

This study had some limitations. Firstly, as we did not include a comparator group of healthy control individuals, it is unknown whether ACC Glx levels were elevated in this CHRp cohort, or whether the effects of diazepam are greater in individuals meeting CHRp criteria compared to in healthy volunteers. However, elevations in ACC Glx in individuals at CHRp is a finding supported by meta-analytical evidence^13,17^. Secondly, despite preclinical evidence implicating the hippocampus, our study lacked measurement of Glx in this region due to significant loss of data due to data quality issues. Nonetheless, more evidence suggests that the ACC is the primary site of Glx changes in CHRp individuals. Although our primary analysis found significant reductions of ACC Glx under diazepam, our study was underpowered (post-hoc power of primary analysis = 0.60). While our study serves as preliminary evidence of diazepam’s Glx modulation efficacy, to address these two limitations, future studies should aim to replicate findings with larger sample sizes. Lastly, we did not measure diazepam plasma levels, preventing analysis of Glx changes relative to achieved drug dose. Future studies should incorporate blood sampling post-MRI to address this.

Overall, this study provides preliminary evidence that a single dose of the benzodiazepine diazepam reduces ACC Glx levels compared to placebo in CHRp individuals. This finding suggests a novel therapeutic mechanism which may be of benefit in individuals with psychosis vulnerability. The development of more selective GABA-enhancing compounds could allow similar regulation of brain excitation in people with psychosis risk while avoiding some of the unwanted side effects of less-selective benzodiazepines.

## Disclosures & Funding

AE has received consultancy fees from Leal Therapeutics. AAG has received consulting fees from Alkermes, Lundbeck, Takeda, Roche, Lyra, Concert and research funding from Lundbeck, Newron and Merck. AAG has received consulting fees from Alkermes, Lundbeck, Takeda, Roche, Lyra, Concert, Newron and SynAgile, and research funding from Newron and Merck. SCRW has recently received research funding from Boehringer Ingelheim and GE Healthcare to perform investigator-led research.

This research was funded by the Wellcome Trust and The Royal Society (202397/Z/16/Z to GM) and the National Institute for Health and Care Research (NIHR) Maudsley Biomedical Research Centre (BRC). The views expressed are those of the authors and not necessarily those of the Welcome Trust, NIHR or the Department of Health and Social Care. For the purpose of open access, the author has applied a CC-BY public copyright licence to any Author Accepted Manuscript version arising from this submission.

## Data Availability

Data produced in the present study are available upon reasonable request to the authors

## Notes

### Clinical Trial

NCT06190483

### Author Declarations

The study received ethical approval from the National Health Service UK Research Ethics Committee (18/LO/0618) and each participant gave written informed consent. While the study received ethical clearance as not a Clinical Trial of an Investigational Medicinal Product by the EU directive 2001/20/EC it was registered on clinicaltrials.gov (NCT06190483)

## References

1. Salazar de Pablo G, Radua J, Pereira J, et al. Probability of Transition to Psychosis in Individuals at Clinical High Risk: An Updated Meta-analysis. JAMA Psychiatry. 2021;78(9):970–978. doi:10.1001/jamapsychiatry.2021.0830

2. Davies C, Cipriani A, Ioannidis JPA, et al. Lack of evidence to favor specific preventive interventions in psychosis: a network meta-analysis. World Psychiatry. 2018;17(2):196–209. doi:10.1002/wps.20526

3. Fusar-Poli P, Estradé A, Stanghellini G, et al. The lived experience of psychosis: a bottom-up review co-written by experts by experience and academics. World Psychiatry. 2022;21(2):168–188. doi:10.1002/wps.20959

4. Benes FM, Berretta S. GABAergic Interneurons: Implications for Understanding Schizophrenia and Bipolar Disorder. Neuropsychopharmacology. 2001;25(1):1–27. doi:10.1016/S0893-133X(01)00225-1

5. Lewis DA, Hashimoto T, Volk DW. Cortical Inhibitory Neurons and Schizophrenia. Nature Reviews Neuroscience. 2005;6:312–324. doi:10.1038/nrn1648

6. Konradi C, Yang CK, Zimmerman EI, et al. Hippocampal interneurons are abnormal in schizophrenia. Schizophrenia Research. 2011;131(1-3):165–173. doi:10.1016/j.schres.2011.06.007

7. Benes FM, Lim B, Matzilevich D, Walsh JP, Subburaju S, Minns M. Regulation of the GABA cell phenotype in hippocampus of schizophrenics and bipolars. Proceedings of the National Academy of Sciences. 2007;104(24):10164–10169. doi:10.1073/pnas.0703806104

8. Charych EI, Liu F, Moss SJ, Brandon NJ. GABA_A_ receptors and their associated proteins: Implications in the etiology and treatment of schizophrenia and related disorders. Neuropharmacology. 2009;57(5-6):481–495. doi:10.1016/j.neuropharm.2009.07.027

9. Egerton A, Modinos G, Ferrera D, McGuire PK. Neuroimaging studies of GABA in schizophrenia: a systematic review with meta-analysis. Translational Psychiatry. 2017;7(6):e1147–e1147. doi:10.1038/tp.2017.124

10. Marques TR, Ashok AH, Angelescu I, et al. GABA-A receptor differences in schizophrenia: a positron emission tomography study using [11C]Ro154513. Molecular Psychiatry. 2021;26:2616–2625. doi:10.1038/s41380-020-0711-y

11. Frankle WG, Cho RY, Prasad KM, et al. In vivo measurement of GABA transmission in healthy subjects and schizophrenia patients. Am J Psychiatry. 2015;172(11):1148–1159. doi:10.1176/appi.ajp.2015.14081031

12. Kang JI, Park HJ, Kim SJ, et al. Reduced Binding Potential of GABA-A/Benzodiazepine Receptors in Individuals at Ultra-high Risk for Psychosis: An [18F]-Fluoroflumazenil Positron Emission Tomography Study. Schizophr Bull. 2014;40(3):548–557. doi:10.1093/schbul/sbt052

13. Merritt K, Egerton A, Kempton MJ, Taylor MJ, McGuire PK. Nature of glutamate alterations in schizophrenia a meta-analysis of proton magnetic resonance spectroscopy studies. JAMA Psychiatry. Published online 2016. doi:10.1001/jamapsychiatry.2016.0442

14. Nakahara T, Tsugawa S, Noda Y, et al. Glutamatergic and GABAergic metabolite levels in schizophrenia-spectrum disorders: a meta-analysis of 1H-magnetic resonance spectroscopy studies. Molecular Psychiatry. 2022;27:744–757. doi:10.1038/s41380-021-01297-6

15. Kraguljac N V., White DM, Reid MA, Lahti AC. Increased hippocampal glutamate and volumetric deficits in unmedicated patients with schizophrenia. JAMA Psychiatry. 2013;70(12):1294–1302. doi:10.1001/jamapsychiatry.2013.2437

16. Bossong MG, Antoniades M, Azis M, et al. Association of Hippocampal Glutamate Levels with Adverse Outcomes in Individuals at Clinical High Risk for Psychosis. JAMA Psychiatry. 2019;76(2):199–207. doi:10.1001/jamapsychiatry.2018.3252

17. Wang Y ming, Xiao Y hui, Xie W lan. Metabolite abnormalities in psychosis risk: A meta-analysis of proton magnetic resonance spectroscopy studies. Asian Journal of Psychiatry. 2020;54:102220. doi:10.1016/j.ajp.2020.102220

18. Merritt K, McGuire PK, Egerton A, 1H-MRS in Schizophrenia Investigators. Association of Age, Antipsychotic Medication, and Symptom Severity in Schizophrenia With Proton Magnetic Resonance Spectroscopy Brain Glutamate Level: A Mega-analysis of Individual Participant-Level Data. JAMA Psychiatry. 2021;78(6):667–681. doi:10.1001/jamapsychiatry.2021.0380

19. Grace AA, Gomes F V. The Circuitry of Dopamine System Regulation and its Disruption in Schizophrenia: Insights Into Treatment and Prevention. Schizophrenia Bulletin. 2019;45(1):148–157. doi:10.1093/schbul/sbx199

20. Gomes FV, Zhu X, Grace AA. Stress during critical periods of development and risk for schizophrenia. Schizophrenia Research. 2019;213:107–113. doi:10.1016/j.schres.2019.01.030

21. Lodge DJ, Grace AA. Aberrant hippocampal activity underlies the dopamine dysregulation in an animal model of schizophrenia. Journal of Neuroscience. 2007;27(42):11424–11430. doi:10.1523/JNEUROSCI.2847-07.2007

22. Du Y, Grace AA. Peripubertal Diazepam Administration Prevents the Emergence of Dopamine System Hyperresponsivity in the MAM Developmental Disruption Model of Schizophrenia. Neuropsychopharmacology. 2013;38(10):1881–1888. doi:10.1038/npp.2013.101

23. Du Y, Grace AA, Hall L. Loss of Parvalbumin in the Hippocampus of MAM Schizophrenia Model Rats Is Attenuated by Peripubertal Diazepam. International Journal of Neuropsychopharmacology. 2016;19(11):1–5. doi:10.1093/ijnp/pyw065

24. Du Y, Grace AA, Hall L. Loss of Parvalbumin in the Hippocampus of MAM Schizophrenia Model Rats Is Attenuated by Peripubertal Diazepam. International Journal of Neuropsychopharmacology. 2016;19(11):1–5. doi:10.1093/ijnp/pyw065

25. Del Pino I, Garcia-Frigola C, Dehorter N, et al. Erbb4 deletion from fast-spiking interneurons causes schizophrenia-like phenotypes. Neuron. 2013;79(6):1152–1168. doi:10.1016/j.neuron.2013.07.010

26. Wen L, Lu YS, Zhu XH, et al. Neuregulin 1 regulates pyramidal neuron activity via ErbB4 in parvalbumin-positive interneurons. PNAS. 2010;107(3):1211–1216. doi:10.1073/pnas.0910302107

27. Kiemes A, Serrano Navacerrada ME, Kim E, et al. Erbb4 Deletion From Inhibitory Interneurons Causes Psychosis-Relevant Neuroimaging Phenotypes. Schizophrenia Bulletin. Published online December 27, 2022:sbac192. doi:10.1093/schbul/sbac192

28. Fusar-Poli P, Spencer T, De Micheli A, Curzi V, Nandha S, McGuire P. Outreach and support in South-London (OASIS) 2001—2020: Twenty years of early detection, prognosis and preventive care for young people at risk of psychosis. European Neuropsychopharmacology. 2020;39:111–122. doi:10.1016/j.euroneuro.2020.08.002

29. Stone JM, Day F, Tsagaraki H, et al. Glutamate Dysfunction in People with Prodromal Symptoms of Psychosis: Relationship to Gray Matter Volume. Biol Psychiatry. 2009;66:533–539. doi:10.1016/j.biopsych.2009.05.006

30. Faul F, Erdfelder E, Buchner A, Lang AG. Statistical power analyses using G*Power 3.1: Tests for correlation and regression analyses. Behaviour Research Methods. 2009;41(4):1149–1160. doi:10.3758/BRM.41.4.1149

31. Yung AR, Yuen HP, Mcgorry PD, et al. Mapping the onset of psychosis: the Comprehensive Assessment of At-Risk Mental States. Australian and New Zealand Journal of Psychiatry Research Assistant. 2005;39:964–971. doi:10.1080/j.1440-1614.2005.01714.x

32. Velthorst E, Levine SZ, Henquet C, et al. To cut a short test even shorter: Reliability and validity of a brief assessment of intellectual ability in Schizophrenia - A control-case family study. Cognitive Neuropsychiatry. 2013;18(6):574–593. doi:10.1080/13546805.2012.731390

33. Zlotnik A, Gruenbaum BF, Mohar B, et al. The Effects of Estrogen and Progesterone on Blood Glutamate Levels: Evidence from Changes of Blood Glutamate Levels During the Menstrual Cycle in Women. Biology of Reproduction. 2011;84(3):581–586. doi:10.1095/biolreprod.110.088120

34. Epperson CN, Haga K, Mason GF, et al. Cortical γ-Aminobutyric Acid Levels Across the Menstrual Cycle in Healthy Women and Those With Premenstrual Dysphoric Disorder: A Proton Magnetic Resonance Spectroscopy Study. Archives of General Psychiatry. 2002;59(9):851–858. doi:10.1001/archpsyc.59.9.851

35. Hamilton M. The assessment of anxiety states by rating. British Journal of Medical Psychology. 1959;32(1):50–55. doi:10.1111/j.2044-8341.1959.tb00467.x

36. Hamilton M. A rating scale for depression. J Neurol Neurosurg Psychiatry. 1960;23(1):56–62. doi:10.1136/jnnp.23.1.56

37. Cornblatt BA, Auther AM, Niendam T, et al. Preliminary findings for two new measures of social and role functioning in the prodromal phase of schizophrenia. Schizophrenia Bulletin. 2007;33(3):688–702. doi:10.1093/schbul/sbm029

38. Greenblatt DJ, Allen MD, MacLaughlin DS, Harmatz JS, Shader RI. Diazepam absorption: Effect of antacids and food. Clinical Pharmacology and Therapeutics. 1978;24(5):600–609. doi:10.1002/cpt1978245600

39. Valium (Diazepam). Hoffman-La Roche, Inc.; 2016. https://www.accessdata.fda.gov/drugsatfda_docs/label/2016/013263s094lbl.pdf

40. Friedman H, Greenblatt DJ, Peters GR, et al. Pharmacokinetics and pharmacodynamics of oral diazepam: Effect of dose, plasma concentration, and time. Clinical Pharmacology & Therapeutics. 1992;52(2):139–150. doi:10.1038/clpt.1992.123

41. Williams EJ. Experimental Designs Balanced for the Estimation of Residual Effects of Treatments. Aust J Chem. 1949;2(2):149–168. doi:10.1071/ch9490149

42. Egerton A, Stone JM, Chaddock CA, et al. Relationship Between Brain Glutamate Levels and Clinical Outcome in Individuals at Ultra High Risk of Psychosis. Neuropsychopharmacology. 2014;39:2891–2899. doi:10.1038/npp.2014.143

43. Truong V, Duncan NW. Suggestions for improving the visualization of magnetic resonance spectroscopy voxels and spectra. R Soc Open Sci. 2020;7(8):200600. doi:10.1098/rsos.200600

44. Abraham A, Pedregosa F, Eickenberg M, et al. Machine learning for neuroimaging with scikit-learn. Frontiers in Neuroinformatics. 2014;8. Accessed March 1, 2023. https://www.frontiersin.org/articles/10.3389/fninf.2014.00014

45. Simpson R, Devenyi GA, Jezzard P, Hennessy TJ, Near J. Advanced processing and simulation of MRS data using the FID appliance (FID-A)-An open source, MATLAB-based toolkit. Magn Reson Med. 2017;77(1):23–33. doi:10.1002/mrm.26091

46. Provencher SW. Estimation of metabolite concentrations from localized in vivo proton NMR spectra. Magnetic Resonance in Medicine. 1993;30(6):672–679. doi:10.1002/mrm.1910300604

47. Provencher S. LCModel and LCMgui User’s Manual - LCModel Version 6.3-1L. Published online October 18, 2016. http://s-provencher.com/lcm-manual.shtml

48. Lin A, Andronesi O, Bogner W, et al. Minimum Reporting Standards for in vivo Magnetic Resonance Spectroscopy (MRSinMRS): Experts’ consensus recommendations. NMR in Biomedicine. 2021;34(5):e4484. doi:10.1002/nbm.4484

49. Musazzi L, Treccani G, Mallei A, Popoli M. The Action of Antidepressants on the Glutamate System: Regulation of Glutamate Release and Glutamate Receptors. Biological Psychiatry. 2013;73(12):1180–1188. doi:10.1016/j.biopsych.2012.11.009

50. Durazzo TC, Meyerhoff DJ, Mon A, Abé C, Gazdzinski S, Murray DE. Chronic Cigarette Smoking in Healthy Middle-Aged Individuals Is Associated With Decreased Regional Brain N-acetylaspartate and Glutamate Levels. Biological Psychiatry. 2016;79(6):481–488. doi:10.1016/j.biopsych.2015.03.029

51. O’Neill J, Diaz MP, Alger JR, et al. Smoking, tobacco dependence, and neurometabolites in the dorsal anterior cingulate cortex. Mol Psychiatry. 2023;28(11):4756–4765. doi:10.1038/s41380-023-02247-0

52. O’Neill J, Tobias MC, Hudkins M, et al. Thalamic glutamate decreases with cigarette smoking. Psychopharmacology (Berl). 2014;231(13):2717–2724. doi:10.1007/s00213-014-3441-5

53. Hädel S, Wirth C, Rapp M, Gallinat J, Schubert F. Effects of age and sex on the concentrations of glutamate and glutamine in the human brain. Journal of Magnetic Resonance Imaging. 2013;38(6):1480–1487. doi:10.1002/jmri.24123

54. Maková M, Kašparová S, Tvrdík T, et al. Mirtazapine modulates Glutamate and GABA levels in the animal model of maternal depression. MRI and 1H MRS study in female rats. Behavioural Brain Research. 2023;442:114296. doi:10.1016/j.bbr.2023.114296

55. Danbolt NC. Glutamate uptake. Progress in Neurobiology. 2001;65(1):1–105. doi:10.1016/S0301-0082(00)00067-8

56. Tani H, Dulla CG, Farzampour Z, Taylor-Weiner A, Huguenard JR, Reimer RJ. A local glutamate-glutamine cycle sustains synaptic excitatory transmitter release. Neuron. 2014;81(4):888–900. doi:10.1016/j.neuron.2013.12.026

57. Snyder J, Wilman A. Field strength dependence of PRESS timings for simultaneous detection of glutamate and glutamine from 1.5 to 7T. J Magn Reson. 2010;203(1):66–72. doi:10.1016/j.jmr.2009.12.002

58. Rothman DL, Feyter HMD, de Graaf RA, Mason GF, Behar KL. 13C MRS studies of neuroenergetics and neurotransmitter cycling in humans. NMR in biomedicine. 2011;24(8):943. doi:10.1002/nbm.1772

59. Rothman DL, Sibson NR, Hyder F, Shen J, Behar KL, Shulman RG. In vivo nuclear magnetic resonance spectroscopy studies of the relationship between the glutamate-glutamine neurotransmitter cycle and functional neuroenergetics. Philos Trans R Soc Lond B Biol Sci. 1999;354(1387):1165–1177. doi:10.1098/rstb.1999.0472

60. Livingston NR, Kiemes A, Devenyi GA, et al. Effects of diazepam on hippocampal blood flow in people at clinical high risk for psychosis. Neuropsychopharmacol. 2024;49(9):1448–1458. doi:10.1038/s41386-024-01864-9

61. Jacob TC, Michels G, Silayeva L, Haydon J, Succol F, Moss SJ. Benzodiazepine treatment induces subtype-specific changes in GABA_A_ receptor trafficking and decreases synaptic inhibition. Proceedings of the National Academy of Sciences. 2012;109(45):18595–18600. doi:10.1073/pnas.1204994109

62. Engin E, Benham RS, Rudolph U. An Emerging Circuit Pharmacology of GABA(A) Receptors. Trends Pharmacol Sci. 2018;39(8):710–732. doi:10.1016/j.tips.2018.04.003

63. Gill KM, Lodge DJ, Cook JM, Aras S, Grace AA. A novel α5GABA(A)R-positive allosteric modulator reverses hyperactivation of the dopamine system in the MAM model of schizophrenia. Neuropsychopharmacology. 2011;36(9):1903–1911. doi:10.1038/npp.2011.76

64. Donegan JJ, Boley AM, Yamaguchi J, Toney GM, Lodge DJ. Modulation of extrasynaptic GABA(A) alpha 5 receptors in the ventral hippocampus normalizes physiological and behavioral deficits in a circuit specific manner. Nat Commun. 2019;10(1):2819. doi:10.1038/s41467-019-10800-1

65. Merritt K, McCutcheon RA, Aleman A, et al. Variability and magnitude of brain glutamate levels in schizophrenia: a meta and mega-analysis. Mol Psychiatry. Published online February 17, 2023:1–10. doi:10.1038/s41380-023-01991-7

66. Markram H, Toledo-Rodriguez M, Wang Y, Gupta A, Silberberg G, Wu C. Interneurons of the neocortical inhibitory system. Nat Rev Neurosci. 2004;5(10):793–807. doi:10.1038/nrn1519

67. Porges EC, Jensen G, Foster B, Edden RA, Puts NA. The trajectory of cortical GABA across the lifespan, an individual participant data meta-analysis of edited MRS studies. Baker CI, Clarke W, eds. eLife. 2021;10:e62575. doi:10.7554/eLife.62575

68. Jiménez-Balado J, Eich TS. GABAergic dysfunction, neural network hyperactivity and memory impairments in human aging and Alzheimer’s disease. Semin Cell Dev Biol. 2021;116:146–159. doi:10.1016/j.semcdb.2021.01.005

69. Sigel E, Steinmann ME. Structure, Function, and Modulation of GABA A Receptors. The Journal of Biological Chemistry. 2012;287(48):40224–40231. doi:10.1074/jbc.R112.386664

