## Supplementary Materials for "Diazepam modulates anterior cingulate glutamate levels in people at clinical high-risk for psychosis"

**Supplementary Methods**

**Table S1.** MRSinMRS checklist

| | |
|---|---|
| 1. Hardware | |
| a. Field strength [T] | 3 T |
| b. Manufacturer | General Electric |
| c. Model (software version if available) | MR750 |
| d. RF coils: nuclei (transmit/receive), number of channels, type, body part | 32-channel head coil |
| e. Additional hardware | N/A |
| 2. Acquisition | |
| a. Pulse sequence | Point-resolved spectroscopy (PRESS) |
| b. Volume of Interest (VOI) locations | Anterior cingulate cortex |
| c. Nominal VOI size [cm$^3$, mm$^3$] | 2 x 2 x 2 cm$^3$ |
| d. Repetition Time (TR), Echo Time (TE) [ms, s] | TR 3000 ms, TE 30 ms |
| e. Total number of Excitations or acquisitions per spectrum<br>In time series for kinetic studies<br>i. Number of Averaged spectra (NA) per time-point<br>ii. Averaging method (e.g. block-wise or moving average)<br>iii. Total number of spectra (acquired / in time-series) | 96 acquisitions with an 8-step phase cycle. For the unsuppressed water, 16 acquisitions with an 8-step phase cycle. |
| f. Additional sequence parameters (spectral width in Hz, number of spectral points, frequency offsets)<br>If STEAM:, Mixing Time (TM)<br>If MRSI: 2D or 3D, FOV in all directions, matrix size, acceleration factors, sampling method | 5000 Hz, 4096 points, -2 ppm frequency offset |
| g. Water Suppression Method | Chemically selective suppression (CHESS) |
| h. Shimming Method, reference peak, and thresholds for “acceptance of shim” chosen | Automated B0 field mapping to < 7 Hz |
| i. Triggering or motion correction method<br>(respiratory, peripheral, cardiac triggering, incl. device used and delays) | N/A |
| 3. Data analysis methods and outputs | |
| a. Analysis software | FID-A, LCModel 6.3-1N |

| | |
|---|---|
| b. Processing steps deviating from quoted reference or product | None |
| c. Output measure<br><br>(e.g. absolute concentration, institutional units, ratio) | Water-scaled (Estimated mM) |
| d. Quantification references and assumptions, fitting model assumptions | Default basis set provided in LCModel 6.3-1N (press_te30_3t_v3.basis) of 17 metabolites (L-alanine, aspartate, creatine (Cr), phosphocreatine (PCr), γ-aminobutyric acid, glucose, glutamine, glutamate (Glu), glutathione, glycero-phosphocholine, phosphocholine, myo-Inositol (mI), L-lactate, N-acetylaspartate (NAA), N-acetylaspartylglutamate, scyllo-inositol, and taurine) |
| **4. Data Quality** | |
| a. Reported variables<br><br>(SNR, Linewidth (with reference peaks)) | S/N: Placebo 33.46 ± 4.92; Diazepam 33.19 ± 6.84<br><br>FWHM (ppm as reported by LCModel): Placebo 0.03 ± 0.01; Diazepam 0.03 ± 0.01 |
| b. Data exclusion criteria | FWHM>2SD; S/N<2SD; CRLB>20%; visual inspection |
| c. Quality measures of postprocessing Model fitting (e.g. CRLB, goodness of fit, SD of residual) | CRLB of Glx: Placebo 5.39 ± 0.68; Diazepam 5.66 ± 0.67 |
| d. Sample Spectrum | Figure 1; Figure S1 |

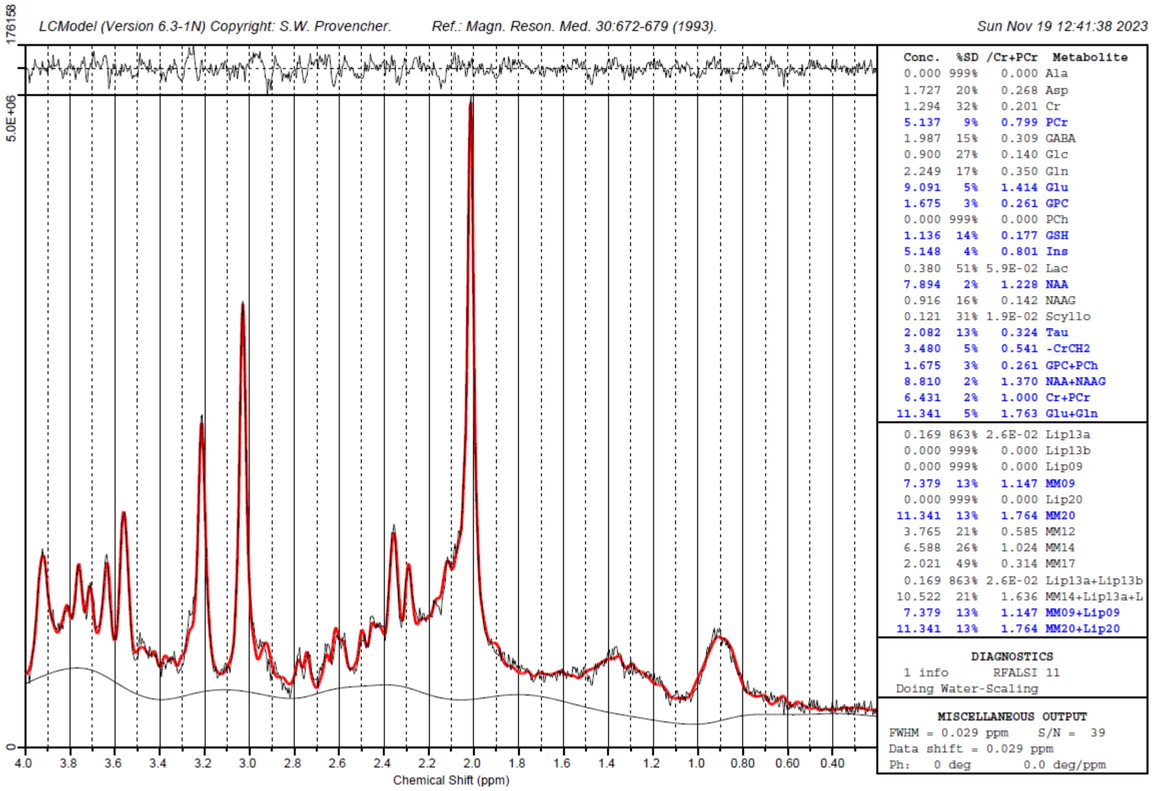


**Figure S1. Sample $^1$H-MRS spectrum**

*CAARMS composite scores*

CAARMS positive composite scores were calculated by multiplying the severity and the frequency of each positive domain subscale (unusual thought content, non-bizarre ideas, perceptual abnormalities, disorganized speech) and summing the products together. Similarly, the CAARMS negative composite score was ascertained using this method on the negative domain subscales (alogia, avolition/apathy, anhedonia).

*Sensitivity Analysis*

Our main analysis of Glx change between diazepam and placebo conditions used median scores from bootstrapped linear combination modelling. A sensitivity analysis was performed using mean value from replicate data fitting.

**Supplementary Results**

*Main Analysis*

**Table S2.** Mixed effects model covariate statistics.

| | t-value | df | p-value | d |
|---|---|---|---|---|
| **First supplementary model** | | | | |
| *Treatment* | -2.110 | 20.749 | 0.047 | 0.46 |
| *Scanning order* | 0.565 | 19.837 | 0.579 | 0.13 |
| *Interscan interval* | 0.010 | 19.563 | 0.992 | 0.002 |
| **Second supplementary model** | | | | |
| *Treatment* | -2.037 | 20.857 | 0.055 | 0.45 |
| *Scanning order* | 0.403 | 16.514 | 0.692 | 0.10 |
| *Interscan interval* | 0.067 | 16.355 | 0.948 | 0.02 |
| *Age* | 0.620 | 16.858 | 0.544 | 0.15 |
| *Sex* | -0.866 | 16.549 | 0.399 | 0.21 |
| *Antidepressant treatment status* | 0.652 | 16.149 | 0.524 | 0.16 |
| *Cigarette use* | -0.633 | 15.713 | 0.536 | 0.16 |

*Sensitivity Analysis*

Linear mixed model using mean values from replicate data fitting analysis showed a significant effect of treatment condition, $t(20.9) = -2.16$, $p = 0.04$, d = 0.47 (Figure S2). Similarly to our main analysis, when accounting for treatment order, interscan interval and data quality, we found also found a significant effect of treatment condition, $t(20.8) = -2.14$, $p = 0.04$, d = 0.47. Lastly, when controlling for further covariates of no interest (age and sex), effect of treatment became non-significant ($t(20.8) = -2.08$, $p = 0.051$, d = 0.46).

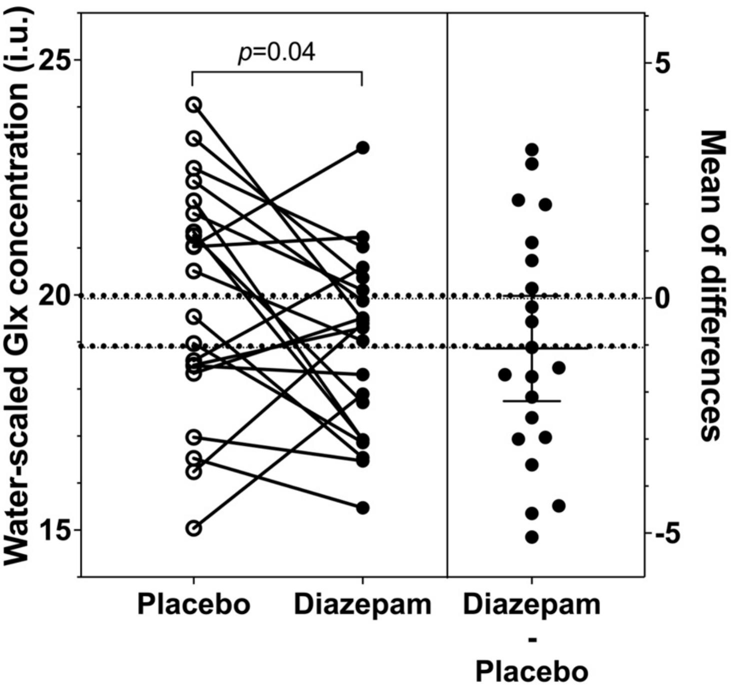


**Figure S2. Glx levels in the anterior cingulate cortex (sensitivity analysis using means from replicate linear combination modelling).** Individual Glx levels during placebo and diazepam are presented on the left and individual diazepam-induced Glx change (diazepam – placebo) with mean and 95% CI are presented on the right. Incomplete datasets were removed for illustrative purposes. Glx, glutamate + glutamine.

**Table S3.** Regressors in multiple regression models.

| | **t-value** | **df** | **p-value** | **d** |
|---|---|---|---|---|
| **Model 1** | | | | |
| *CAARMS positive composite score* | 0.476 | 16 | 0.641 | 0.24 |
| *CAARMS negative composite score* | -0.865 | 16 | 0.579 | 0.43 |
| **Model 2** | | | | |
| *CAARMS positive composite score* | 0.144 | 12 | 0.888 | 0.08 |
| *CAARMS negative composite score* | 1.476 | 12 | 0.166 | 0.85 |
| *Age* | -4.360 | 12 | 0.001 | 2.52 |
| *Sex* | 0.616 | 12 | 0.549 | 0.36 |
| *Cigarettes per day* | -0.672 | 12 | 0.514 | 0.39 |
| *Antidepressant use* | 0.731 | 12 | 0.479 | 0.42 |
